# Unveiling Meropenem Resistance and Co-Resistance Patterns in *Klebsiella pneumoniae* and *Acinetobacter baumannii*: A Global Genome Analysis Using ML/DL and Association Mining

**DOI:** 10.1101/2025.02.04.25321629

**Authors:** Srimathy Ramachandran, K Deepak, M Shrikumaran, R Mohamed Rasiq, S Ananya, V Aruna, P Derrick Daniel, B Sruthi, Mohan S Suma

## Abstract

**Background:** The increasing prevalence of meropenem-resistant gram-negative bacteria has significantly undermined its effectiveness and has increased treatment failure and mortality rates. The global availability of bacterial WGS data with antimicrobial resistance phenotypes enables large-scale genome analysis to explore resistance determinants. This study investigated the meropenem resistance mechanism in multidrug-resistant (MDR) *Klebsiella pneumoniae* (KP) and *Acinetobacter baumannii* (AB) isolates using advanced data analytics approaches.

**Methods:** We analysed 2,411 KP and 375 AB isolates with meropenem-resistant and susceptible phenotypes from the BV-BRC database. AMR genes and mutations were identified from the isolates using the CARD database as a reference. Significant AMR genes and missense mutations, determined through chi-square tests, were subsequently used to train ML and DL models. The best-performing SVM model was used for sequential feature selection to identify key features. Additionally, association mining was conducted separately on the selected features and the antibiotics data.

**Results:** Notable differences were observed in the proportions of genes contributing to the meropenem resistance mechanism categories between KP and AB, including carbapenemases (4% in KP, 23% in AB), antibiotic efflux (30%, 60%), target alteration (23%, 12%), and reduced permeability (18%, 3%). Mutation frequencies also vary, with antibiotic efflux (26%, 67%), target alteration (64%, 5%), and reduced permeability (7%, 15%). A total of 410 significant features in KP and 211 in AB were identified for model building. SVM-based feature selection pinpointed seven key features in KP and 10 in AB, resulting in 95% accuracy for both. Association mining revealed *bla*_KPC-2_, *bla*_KPC-3_, *ble*_MBL_, and *aac(6’)-Ib9* as key factors in KP, and *bla*_OXA-23_, Abau_gyrA_FLO|Ser81Leu, and Abau_OprD_IMP|Asn411Asp in AB associated with meropenem resistance. The observed prevalence of AAC genes and the *gyrA* mutation, along with insights from association mining, reveals the co-resistance of meropenem with aminoglycosides and fluoroquinolones, while *oprD* mutations imply potential shared resistance across antibiotics.

**Conclusion:** The analysis of AMR genes and mutations based on resistance mechanisms revealed distinct differences in meropenem resistance between KP and AB. The ML/DL models and association mining approaches identified key resistance features and cross-antibiotic resistance insights. These findings deepen our understanding of meropenem resistance, enabling more precise and effective antimicrobial interventions.

## INTRODUCTION

Antibiotics have revolutionized medicine by effectively combating infections and supporting important medical procedures. Despite their importance, antibiotics are losing effectiveness as antimicrobial resistance levels rise, presenting a significant challenge to global health (1). Carbapenems are a class of antimicrobial agents specifically designed to treat infections caused by multidrug-resistant microorganisms. They rank as the third most utilised antibiotic in intensive care units (ICUs) worldwide and are the last-resort treatment for critically ill patients because of their broad-spectrum activity against both gram-positive and gram-negative bacteria (2,3). Owing to their structure they are characterized as β-lactam antibiotics, which feature a β-lactam ring fused to a five-membered ring with a carbon atom at position 1, replacing the sulfur found in penicillins, and a double bond between the C-2 and C-3 positions (4). This unique structure provides protection against numerous β-lactamases produced by a wide range of bacterial pathogens.

Like other β-lactam antibiotics, carbapenems inhibit bacterial transpeptidases crucial for synthesizing peptidoglycan within the cell wall by binding to enzymes known as penicillin-binding proteins (PBPs), including PBP1a, 1b, 2, and 3. This prevents bacteria from completing cell wall synthesis, leading to cell death due to autolysin activity (5). Carbapenems enter gram-negative bacteria by passing through outer membrane proteins (OMPs) called porins and through the cell wall in gram-positive bacteria. The overuse of carbapenems has increased carbapenem resistance, allowing bacteria to withstand clinically effective concentrations of carbapenems. Consequently, the prevalence of clinical bacterial isolates producing β-lactamases capable of hydrolysing carbapenems, commonly known as carbapenemases, has increased (6,7). The mechanisms of carbapenem resistance in bacteria involve the synthesis of carbapenemases, alterations in PBPs, overexpression of efflux pumps, and the loss of outer membrane porins (8).

The clinically approved and widely used carbapenems include imipenem, meropenem, ertapenem, and doripenem (9). The binding affinities of carbapenems to different PBPs vary, potentially explaining variations in their antimicrobial effectiveness. Meropenem’s compact size and zwitterionic state facilitate its broad-spectrum effectiveness by enabling easy penetration of the cell membrane in multiple gram-negative bacteria (10). Infections caused by meropenem-resistant bacteria lead to increased treatment failure, longer hospital stays, increased costs, and increased mortality (11).

Whole genome sequencing (WGS) is a powerful tool for understanding antimicrobial resistance (AMR). With increasing amounts of genome data in public repositories, large-scale analysis helps identify resistance markers, track pathogen spread, and uncover resistance mechanisms, aiding in the development of new treatments and public health strategies. By integrating WGS and antimicrobial susceptibility testing, machine learning (ML) and deep learning (DL) methods show great potential in predicting resistance, discovering new resistance genes, and identifying effective treatments (12–14). ML/DL models that utilize feature representations such as k-mers (15–17), pangenomes (18–20) and mutations (17,21–24) have been effectively used to predict AMR phenotypes. There have been efforts to understand antimicrobial resistance mechanisms (14) in different pathogens, including the use of ML and genome-scale metabolic models (GSMs) to investigate links between genetic resistance determinants and metabolism (25). Additionally, metabolic model-based machine learning classifiers suggest a biochemical basis for how the identified alleles contribute to AMR (20).

This study utilized large-scale genome analysis and advanced computational techniques to investigate the mechanisms of resistance to meropenem. Meropenem resistance mechanisms have been studied in region specific manner; however, extensive large-scale genomic analyses to uncover global trends in meropenem resistance determinants are still lacking. The presence and absence of AMR genes, along with missense mutations in these genes, were utilized as features for ML/DL models to explain meropenem resistance. These models were trained on WGS data alongside laboratory-confirmed drug susceptibility data, incorporating data from both meropenem resistant and susceptible strains of two gram-negative bacteria, *Klebsiella pneumoniae* (KP) and *Acinetobacter baumannii* (AB), obtained from the BV-BRC database. Various techniques, including ML/DL approaches, feature selection, and association mining have been employed to identify hidden features contributing to meropenem resistance.

## MATERIALS AND METHODS

### Data collection

The assembled contig files of the KP and AB isolates were sourced from the Bacterial and Viral Bioinformatics Resource Center (BV-BRC) to perform large-scale genomic analysis to explore the role of AMR genes and associated mutations in meropenem resistance. Among the ESKAPE pathogens, KP and AB were found to have the maximum number of genomes available in BV-BRC with WGS raw data and assembled files. We obtained a total of 2,411 isolates of KP and 375 isolates of AB from the BV-BRC database (v3.36.16)(26), on the basis of the following filters: genome status as WGS or complete, genome quality as good, and only SRA ID-available entries. For the AMR phenotype, we filtered by antibiotic (meropenem), laboratory method (broth dilution), and resistance phenotype (resistant and susceptible). For external validation, we retrieved 156 isolates of KP and 107 isolates of AB with experimentally determined meropenem phenotype data from the NCBI Pathogen database (https://www.ncbi.nlm.nih.gov/pathogens/, accessed on 24 September 2024) and BV-BRC.

### AMR gene reference library preparation

An AMR reference library was created using the Comprehensive Antibiotic Resistance Database (CARD)(v3.2.9) (27) to identify AMR genes and mutations in the clinical isolates of KP and AB selected for the study. The FASTA sequences of 5,026 AMR genes, belonging to homology, variant, knockout and overexpression models in the CARD database, were obtained and converted to GenBank format using the Prokka(v1.14.6) which is available on Galaxy platform (28,29). The class details of the beta-lactamase and carbapenemase genes were obtained from the Beta-lactamase database (BLDB) (30) as of 4 January 2025

### Identification of AMR genes and missense mutations and preprocessing

The AMR reference library, comprising 5026 AMR genes, was used to create a custom database for identifying AMR genes using ABRicate v1.0.1. (31). The assembled contigs were then subjected to ABRicate with the custom database, applying a minimum identity and minimum coverage threshold of 90% and the ABRicate summary file was generated. With default parameters, the standalone tool Snippy v4.6.0 (32), was used to identify mutations in AMR genes from the assembled contig files. The Prokka-annotated GenBank file, which is based on the CARD database containing 5026 AMR genes, was utilized as a reference. Using an in-house python script, we extracted missense mutations from the snippy output.vcf file, formatted them, and converted them into a presence-absence matrix. Additionally, we converted the ABRicate summary file into a presence-absence gene matrix and merged both matrices into a single feature table, with samples in the rows and the AMR gene and mutation presence-absence as columns.

### ML/DL model generation, feature selection and evaluation

We employed six ML models and one DL model: Logistic Regression (LR), Decision Tree (DT), Random Forest (RF), Support Vector Machine (SVM), eXtreme Gradient Boosting (XGB), Multi-Layer Perceptron (MLP), and Deep Neural Networks (DNN). The models were implemented using the scikit-learn and keras libraries. Initially, the models were trained using default parameters. Subsequently, six models (LR, DT, RF, SVM, XGB, MLP) were fine-tuned using the BayesSearchCV algorithm with different hyperparameters for each model, along with 5-fold cross-validation. The DNN model was optimized and tuned using the Bayesian optimization algorithm (33), with 5-fold cross-validation. Model evaluation was performed on the basis of accuracy, precision, recall, and F1-score, and the best-performing model was selected. This model was then used for feature selection using the sequential feature selection (SFS) approach from the mlxtend library (34). Using the selected features, the model was trained, tested, and validated using external datasets.

### Association mining and network generation

For the extracted features of both KP and AB, we used the Apriori algorithm from the mlxtend library to identify frequent item sets. A support value of 0.01 was set for KP and 0.06 for AB, ensuring that at least 20 isolates were included in the calculations. We generated association rules with a confidence threshold of 80% or higher. To explore the shared and co-resistance among antibiotics in AB and KP, we analysed antibiotic resistance data available from the isolates used in the present study and identified common resistance patterns using the association rule mining algorithm. The network representation of the association rules was created using the arulesViz package (35) in R (v.4.4.2).

### Phylogenetic analysis

The entire feature table, created on the basis of the presence and absence of AMR genes and missense mutations, was used to calculate the binary distance matrix using an in-house R script. The neighbor-joining method was then applied to construct the phylogenetic tree, which was visualized using iTOL(v7.0) (36).

## RESULTS

### Genomic insights of KP and AB WGS data, which are resistant and susceptible to meropenem

A large-scale global genomic analysis was conducted to investigate the mechanisms of meropenem resistance, using 2,411 KP genomes and 375 AB genomes, sourced from the BV-BRC database. Among the KP isolates, 959 were resistant and 1452 were susceptible to meropenem. For AB, 192 isolates were resistant and 183 were susceptible to meropenem (Table 1). This study profiled the presence of AMR genes and missense mutations in AMR genes, on the basis of the CARD database, to identify the genomic factors contributing to meropenem resistance. A total of 321 AMR genes and 1579 missense mutations in AMR genes were identified in the KP whereas 184 AMR genes and 1119 missense mutations in AMR genes were identified in the AB. Fig. 1 depicts the feature-based phylogeny of all the genomes used in the study (Table S1 and Table S2). In the annotated phylogenetic tree, the resistant and susceptible isolates were mixed within different clades for both organisms (Fig. 1 innermost circle). Country-specific clades are observed in both organisms, with 23 countries represented in KP and eight countries in AB (second innermost circle), and the maximum number of samples observed from the USA in both organisms. The minimum inhibitory concentration (MIC) for meropenem varies across different clades (third circle from inside), with MIC values for KP ranging from 0.12 to 128 mg/mL and for AB ranging from 0.25 to 32 mg/mL. In KP and AB, the majority of the different clades in the phylogeny were found to follow the same sequence type (ST), as shown in the fourth circle from inside Fig. 1a, Fig. 1b and Fig. S1. A total of 215 STs were observed in KP, whereas 75 STs were observed in AB. The isolation years for the collected samples ranged from 2008-2017 for KP and from 2003-2017 for AB, as shown in the second circle from the outside in Fig. 1. The isolates were collected from various sources and were broadly categorized into 10 different groups for both KP and AB, as shown in the outermost circles of Fig. 1a and Fig. 1b. The majority of samples were obtained from the excretion source (986) for KP and the wound source (139) for AB.

**Fig. 1:**
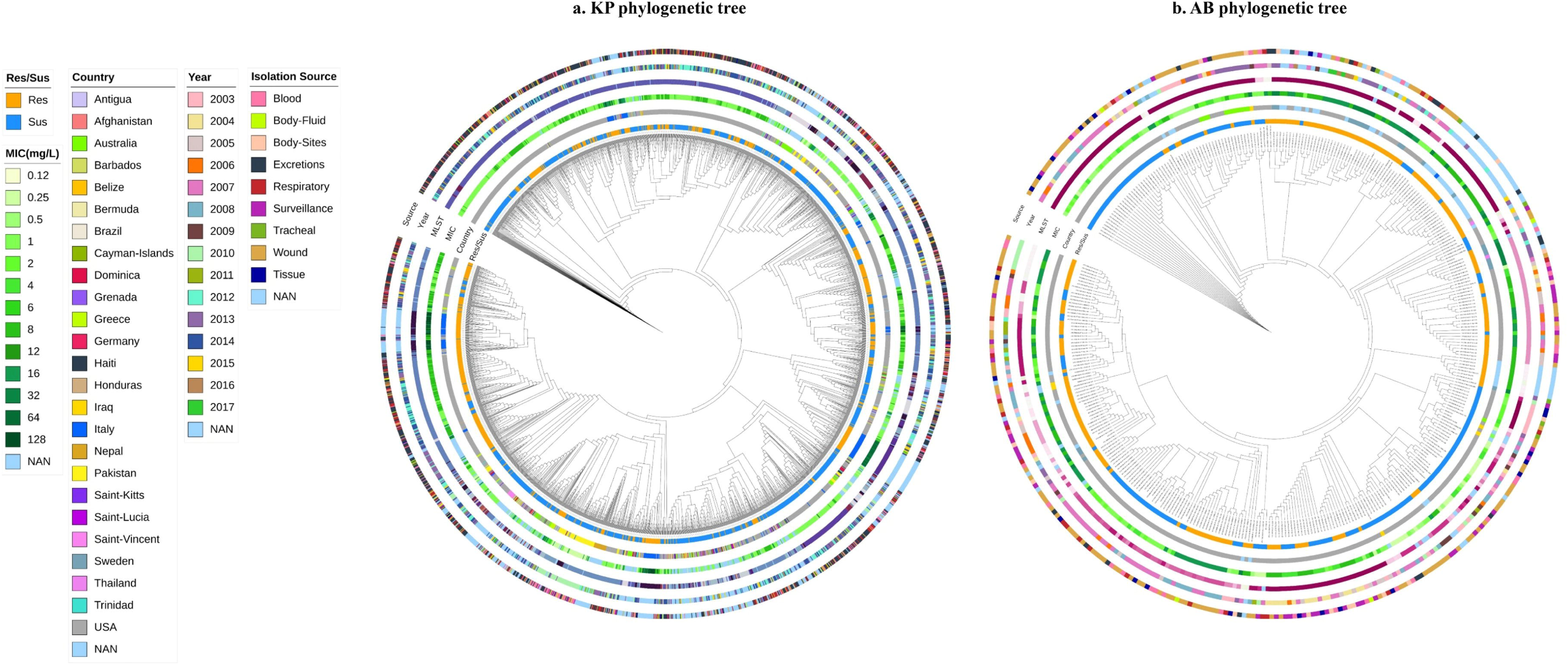
Phylogenetic tree of **a**. *Klebsiella pneumoniae* and **b.** *Acinetobacter baumannii* isolates, were constructed using a presence-absence matrix of AMR genes and missense mutations. The concentric circles represent key metadata from the innermost to the outermost layer: (1) resistant/susceptible phenotype, (2) country of isolation, (3) minimum inhibitory concentration (MIC) values, (4) multi-locus sequence typing (MLST) (shown in Fig. S1), (5) year of isolation, and (6) source of isolation.

**Table 1:**
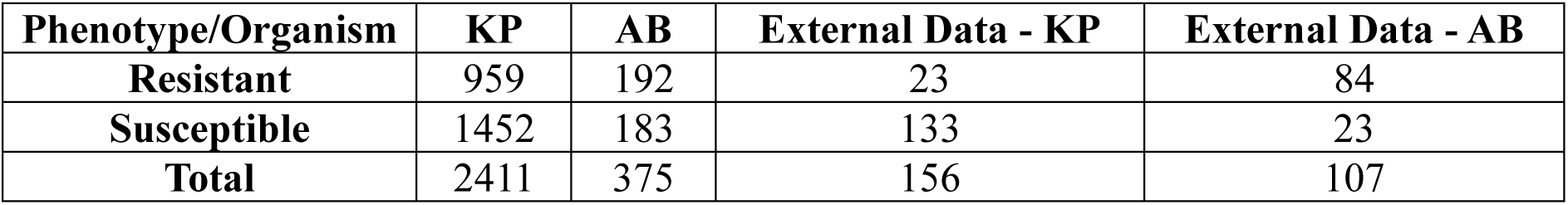
Overall dataset collected for the study.

| Phenotype/Organism | KP | AB | External Data - KP | External Data - AB |
| --- | --- | --- | --- | --- |
| **Resistant** | 959 | 192 | 23 | 84 |
| **Susceptible** | 1452 | 183 | 133 | 23 |
| **Total** | 2411 | 375 | 156 | 107 |

We categorized the AMR genes into six resistance mechanism categories defined by the CARD database: antibiotic inactivation, efflux, target alteration, target protection, target replacement, and reduced permeability to antibiotics (Fig. 2). Antibiotic efflux is the predominant resistance mechanism in both KP and AB, with a greater proportion of efflux genes found in AB (60% of all the AMR genes in AB isolates) than in KP (30%). The proportion of antibiotic inactivation genes in KP was 23% and that in AB was 21%. The proportion of antibiotic target alteration genes was 23% in KP and 12% in AB. Genes associated with reduced permeability to antibiotics are more abundant in KP (18%) than in AB (3%). Antibiotic target replacement genes account for 5% of the genes in KP and 3% of those in AB. Only 1% of the genes in both KP and AB contribute to antibiotic target protection (Fig. 2a, innermost ring). Upon separately analysing the genomes resistant and susceptible to meropenem, we found that the proportions of resistance mechanism classes were similar, with a slight increase observed in the antibiotic inactivation category in the meropenem-resistant phenotype (Fig. 2a and Fig. 2b, middle ring for resistant and outer ring for susceptible). The AMR genes belonging to various AMR mechanism categories are listed in Table S3 and Table S4.

**Fig. 2:**
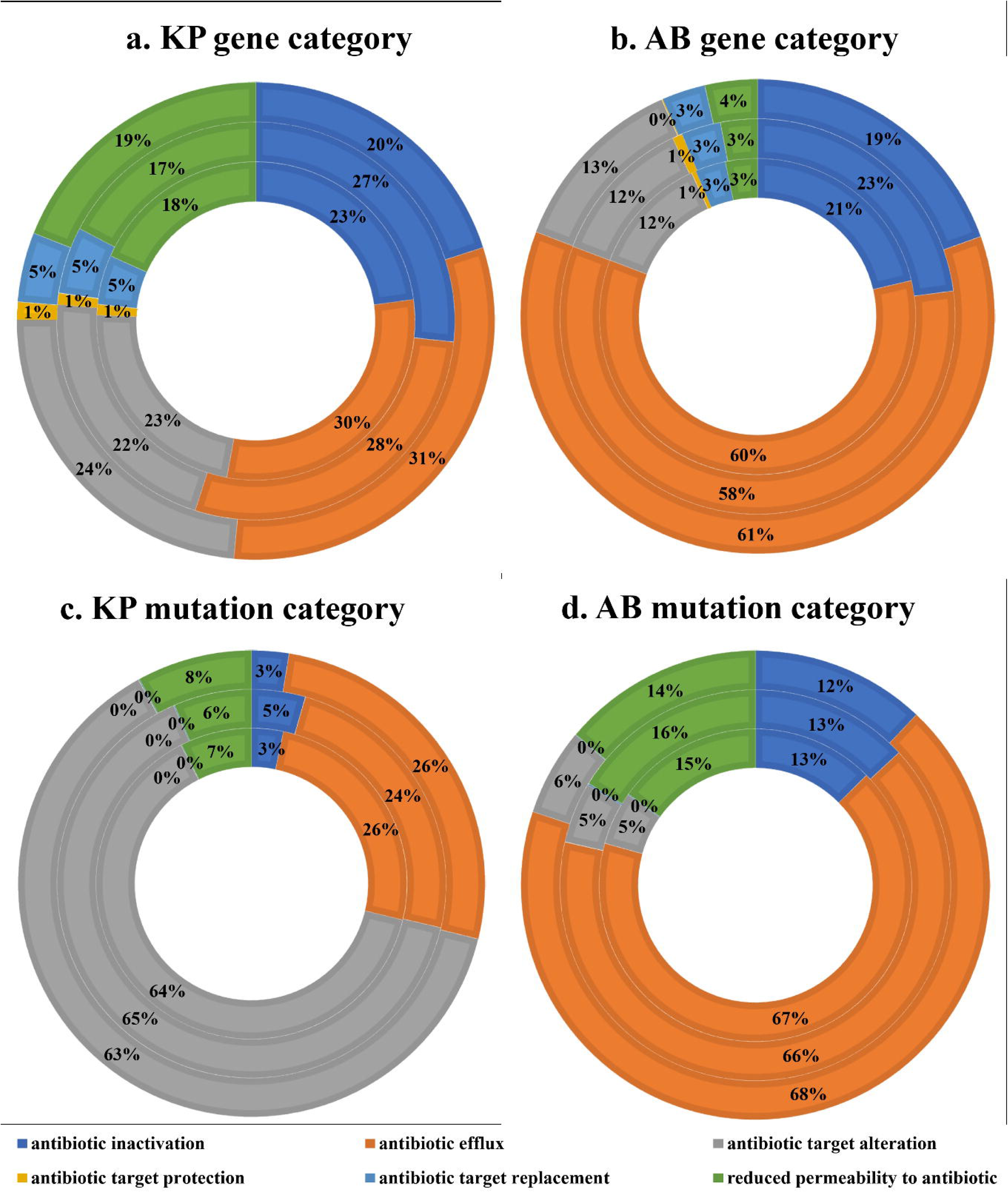
Percentage of AMR genes across different AMR resistance categories based on the CARD database for **a** KP and **b** AB. The circles represent: Circle 1 (innermost) - entire dataset, Circle 2 (middle) - resistant isolates only, and Circle 3 (outermost) - susceptible isolates. Percentage of mutations observed in different AMR resistance categories based on the CARD database for **c** KP and **d** AB. In AB no mutations are observed in the antibiotic protection category and very less mutations are observed in the antibiotic target replacement category.

AMR gene mutation details indicate that approximately 32% of AMR genes undergo mutations in KP, whereas 27% of genes exhibit mutations in AB. The analysis of the mutations revealed that a major difference between organisms was observed in the antibiotic target alteration class, with 64% of mutations in KP and only 5% in AB and the antibiotic efflux category, with 26% of mutations attributed to efflux and corresponding regulator genes in KP and 67% in AB (Fig. 2c and Fig. 2d). Mutations in antibiotic inactivation genes contribute 3% in KP and 13% in AB. The genes associated with reduced permeability to antibiotics accounted for 7% of the mutations in KP and 15% in AB. Antibiotic target protection genes contribute to only 0.01% of mutations in KP and no mutations in AB. The contribution of mutations from antibiotic target replacement genes is 0.07% in KP and 0.08% in AB (Fig. 2c and Fig. 2d innermost ring). The ratio of mutations in the resistant and susceptible categories also reflected the same trend (Fig. 2c and Fig. 2d, middle ring for resistant and outer ring for susceptible). The list of mutated AMR genes belonging to various AMR mechanism categories is listed in Table S5 and Table S6.

To investigate the meropenem resistance mechanism, we focused on key contributing factors, including carbapenemases, efflux pumps, porin genes and PBPs. Carbapenemases, which belong to the antibiotic inactivation category, play a major role in the meropenem resistance mechanism by breaking down carbapenems. Given their importance, we investigated the presence of carbapenemases in the clinical isolates used in our study. Our analysis revealed 16 carbapenemases in KP and 44 in AB on the basis of the BLDB database (highlighted in Table S3 and Table S4). The carbapenemases in KP include Class A enzymes such as *bla*_GES_ and *bla*_KPC_, Class D enzymes such as *bla*_OXA_, and subclass B1 enzymes such as *bla*_IMP_, *bla*_NDM_, and *bla*_VIM_ variants. *bla*_KPC-2_ and *bla*_KPC-3_ are especially prevalent. *bla*_KPC-2_ was found in 51% of the carbapenem-resistant genomes and 2% of the susceptible genomes, whereas *bla*_KPC-3_ was present in 29% of the resistant genomes and in only one genome from the susceptible group (highlighted in Table S3). In AB, subclass B1 carbapenemases such as *bla*_NDM_ and Class D carbapenemases, including *bla*_OXA_ variants, are commonly identified. A total of 43 *bla*_OXA_ variants with carbapenemase activity, along with the *bla*_NDM-1_ variant, have been reported in AB. Among the carbapenemases, *bla*_OXA-23_ and *bla*_OXA-66_ are prevalent in AB. *bla*_OXA-23_ is present in 59% of carbapenem-resistant genomes and 1% of susceptible genomes, whereas *bla*_OXA-66_ is found in 27% of resistant genomes and 33% of susceptible genomes (as highlighted in Table S4). None of the carbapenemases in KP or AB were found to have mutations. We also investigated other beta-lactamase genes, including carbenicillinase, broad-spectrum, narrow-spectrum, and ESBL genes, among the isolates. Among KP, Class A beta-lactamases were the most prevalent, with 113 genes identified, followed by Class C (7) and Class D (6) beta-lactamases. In AB, Class C beta-lactamases were dominant, with 57 genes, followed by Class A (11) and Class D (3) beta-lactamases.

Importantly, non-carbapenemase carbapenem resistance is one of the important observations found in KP isolates reported by Rosas N et al. 2023. We checked the non-carbapenemase isolates in our study. It is observed that 67 (∼7%) of meropenem-resistant KP isolates did not contain any carbapenemase genes. However, 4% of the isolates in the susceptible category possessed carbapenemase. The analysis of non-carbapenemase carbapenem-resistant KP genomes revealed the presence of 117 AMR genes and 215 missense mutations. When analysed other beta-lactamase genes, we identified 27 Class A genes, 3 Class C genes, and 5 Class D genes. More than 90% of the non-carbapenemase isolates harbour genes that contribute to resistance through antibiotic efflux (9 genes), antibiotic target alteration (7 genes), reduced permeability (6 genes), and antibiotic inactivation (1 gene). In terms of mutation frequency, more than 90% of the isolates harboured mutations in antibiotic target alteration genes (7), antibiotic efflux genes (4), and reduced permeability genes (2). In AB, no resistant isolate was found without carbapenemase whereas 98% of the susceptible isolate harbored at least one carbapenemase.

Overall, the analysis of the antibiotic efflux category revealed 50 efflux-associated genes in KP and 27 in AB. In KP, efflux pumps belonging to the RND, SMR, MFS, and ABC families have been identified. The most frequent efflux genes in KP include *acrA* (99.95%), *kpnF* (99.87%), *lptD* (99.71%), and *kpnG* (99.58%), along with efflux regulators such as *acrR* (99.21%) *kpnE* (97.92%), *oqxA* (97.22%), and *oqxB* (96.47%) (Table S3). The efflux genes were found to have multiple mutations and the highly mutated efflux-associated genes in KP include *ramR* (with 91 mutations), *oqxB* (90), *oqxA* (58), and *lptD* (57) as reported in Table S5. The most frequent mutations (>90% of the isolates) are observed in efflux-associated genes such as *acrA*, *kpnG*, and *crp* (Table S5). In AB, efflux pumps belonging to the RND, MATE, SMR, and MFS categories were found. The RND efflux pumps, including *adeI*, *adeJ*, *adeK*, *adeF*, *adeG*, *adeH*, *abuO*, *adeA*, *adeB* along with their regulators, *adeR*, *adeS*, *adeL*, and *adeN*, the MATE family efflux pump *abeM*, the SMR family efflux pump *abeS* and the MFS family efflux pumps *amvA* and *abaF,* are present in more than 90% of the isolates (37) (Table S4). In AB, the ade family genes (*adeA*, *adeB*, *adeC*, *adeJ*, *adeH*, *adeS*, *adeF*) and the *amvA* gene have more missense mutations. Mutations in efflux genes, such as adeJ|Met1043Ile, abeM|Ser233Asn, adeN|ProPhe11HisIle, and the regulator gene adeS|Lys339Gln, were highly prevalent with more than 90% frequency among the isolates (Table S6).

In terms of porins, KP contains 12 porin genes, while AB has 3. In KP, the most frequently found porins included *OmpA* (99.91%), *mdtQ* (99.91%), *OmpK36* (99.41%), *OmpK35* (99.08%), *OmpK37* (98.79%), and *lamB* (96.93%) (Table S3). The *mdtQ* gene in KP is associated with 103 missense mutations. Mutations such as OmpA|Thr332Asn, OmpK37|Ile70Met, and OmpK37|Ile128Met are present in more than 97% of KP isolates (Table S5). In AB, *lpsB* was the most prevalent porin, occurring in 98.66% of the isolates (Table S4). The porin genes *lpsB*, *OprD*, and *carO* in AB harbor multiple missense mutations. Mutations, such as LpsB|Ile157Val, LpsB|Gly146Asp, Abau_OprD_IPM|Asn411Asp, and Abau_OprD_IPM|Thr415_Asn416delinsGly are present in more than 90% of AB isolates (Table S6).

Other AMR mechanism categories in the CARD include antibiotic target alteration, antibiotic target replacement, and antibiotic target protection. In KP, we identified 28 genes in the alteration category, 15 genes in the protection category, and 22 genes in the replacement category. Notably, 7 genes involved in antibiotic target alteration were present in more than 99% of the samples, whereas no genes from the protection or replacement categories showed such prevalence (Table S3). In KP, the genes *phoQ*, *arnT*, *rpoB*, and *eptB* harbor more than 50 different missense mutations (Table S5). Penicillin-binding proteins (PBPs), which are direct targets of carbapenems, fall under antibiotic target alterations according to CARD data. We detected *pbp3* in 99.91% of KP isolates, and it has 40 associated missense mutations, among which PBP-3|Val375Ala was the most frequent (Table S5). For AB, we identified five genes in the alteration category, one gene in the protection category, and six genes in the replacement category. Four of the genes involved in antibiotic alteration were present in more than 98% of the samples, while no genes from the protection or replacement categories showed such prevalence. The *pbp3* gene was absent in AB and the genes *parC* and *gyrA* harbored multiple missense mutations (Table S4 and Table S6).

### ML/DL based identification of features contributing to meropenem resistance

Given the absence of any specific difference in the overall trend of AMR genes and mutation presence between the resistant and susceptible categories in both organisms, an ML/DL based classification approach was used to identify the significant features contributing to meropenem resistance. To identify significant features, we performed a chi-square test (p ≤ 0.05) on the full feature sets of 1900 for KP and 1303 for AB. This resulted in 410 and 211 significant features, respectively, encompassing AMR genes and missense mutations in KP and AB. With these features, we explored six machine learning models (LR, DT, RF, SVM, XGB, and MLP) and DNN in this step and the 5-fold cross-validation accuracy values are reported in Fig. S2. After optimization, the models were evaluated based on key performance metrics, including accuracy, precision, recall, and F1-score. The training and test metric values of all seven models in KP and AB are depicted in Fig. 3a and Fig. 3b, respectively. On the basis of these metrics, the SVM with linear kernel model exhibited the best overall performance for both organisms, achieving test accuracy and F1-score values of 96% and 96% for KP and 93% and 93% for AB, respectively (Fig. 3a and Fig.3b). Subsequently, sequential feature selection was performed using the SVM model to identify the most important features for differentiating resistant and susceptible phenotypes. This process led to the selection of seven features for KP and 10 for AB which can discriminate between resistant and susceptible classes (Fig. 4a and Fig. 4c). When trained and tested using these selected features, the SVM model achieved accuracies of 94% and 92% on the training set (Fig. 4a and Fig.4c) and 95% and 95% on the test datasets of KP and AB respectively (Fig. 4b and Fig. 4d). To assess the model’s generalizability, external validation was performed using the selected features. The SVM model demonstrated robust performance with external validation accuracies of 95% for KP and 94% for AB (Fig. 4b and Fig. 4d).

**Fig. 3:**
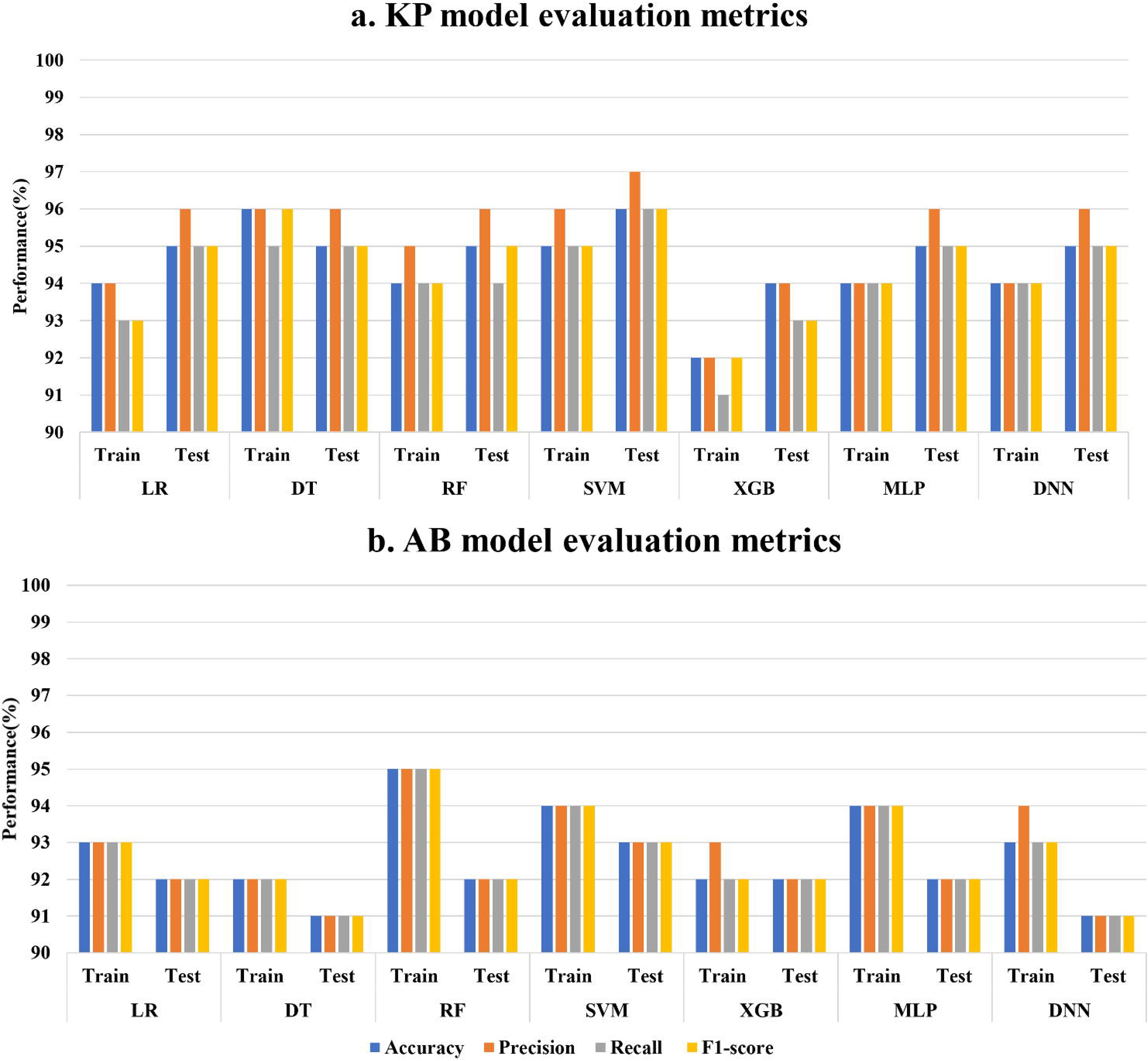
Performance metrics—accuracy, precision, recall, and F1-score—for six ML models and one DL model based on the significant feature set selected based on chi-square test for training and testing datasets of **a** KP and **b** AB

**Fig. 4:**
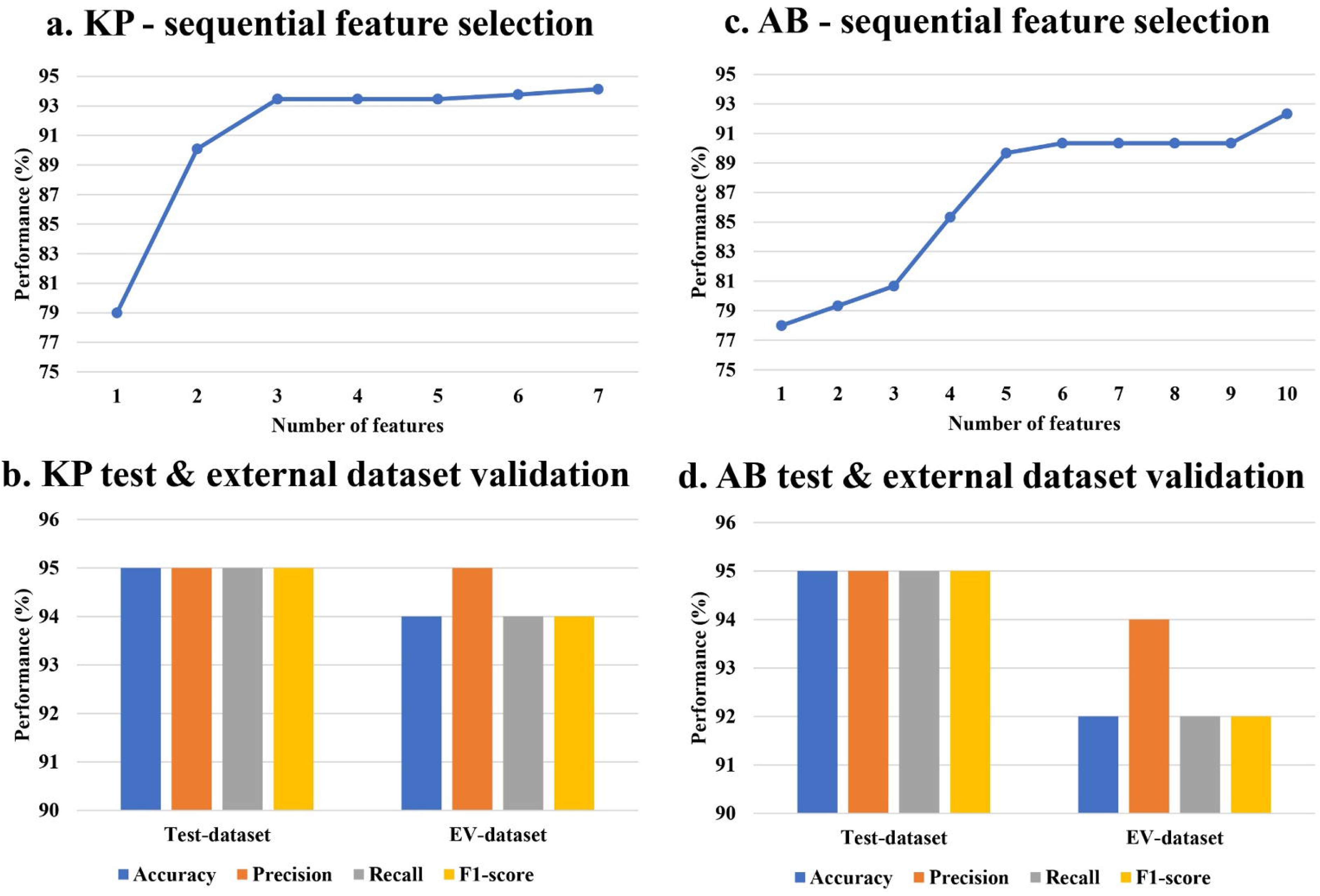
Sequential feature selection using the best-performing SVM model resulted in **a** 7 features for KP reaching 94% training accuracy, and **c** 10 features for AB reaching 92% training accuracy. The test dataset and the external dataset validation performed using the selected features yielded 95% & 94% accuracy in **b** KP and 95% & 92% accuracy in **d** AB

The seven selected key features of the KP are reported in Table 2. Among the AMR genes, carbapenemases *bla*_KPC-2_, and *bla*_KPC-3_, metallo-β-lactamase *ble*_MBL_ and aminoglycoside acetyltransferase *aac(6’)-Ib9* were identified as key features. Three mutations were identified in genes such as *oqxA*, *kpnE* and *kpnG*. The AMR genes, *bla*_KPC-2_, *bla*_KPC-3_, *ble*_MBL_, and *aac(6’)-Ib* were significant and were found predominantly in the resistant phenotype category whereas the mutations oqxA|Lys308Gln, Kpne_KpnE|Ile96Val, and Kpne_KpnG|Ala69Ser were found significantly in genomes with meropenem susceptible phenotype (Table 2).

**Table 2:** Details of seven features identified from SVM-based feature selection for KP.

| KP feature name | SVM-coefficient | Chi-square p-value | Resistant (%) | Susceptible (%) |
| --- | --- | --- | --- | --- |
| *oqxA*\|Lys308Gln | -1.89 | 1.74E-06 | 7(0.73%) | 59(4.06%) |
| *Kpne_KpnE*\|Ile96Val | -1.89 | 6.19E-08 | 7(0.73%) | 69(4.75%) |
| *Kpne_KpnG*\|Ala69Ser | 1.89 | 8.99E-06 | 18(1.88%) | 82(5.65%) |
| *ble*<sub>MBL</sub> | 1.999 | 2.9E-23 | 91(9.49%) | 14(0.96%) |
| *bla*<sub>KPC-2</sub> | 1.999 | 7.2E-180 | 490(51.09%) | 30(2.07%) |
| *bla*<sub>KPC-3</sub> | 2 | 3.3E-103 | 276(28.78%) | 1(0.07%) |
| *aac*(6')-Ib9 | 1.999 | 3.44E-14 | 47(4.90%) | 4(0.28%) |

Table 3 lists the 10 key features identified for AB. The six AMR genes, *bla*_OXA-23_, *bla*_OXA-113_, *bla*_OXA-174_, *bla*_OXA-312_, *bla*_OXA-72_, and *aac(6’)-Ib7*, were identified as key features showing a significant count with the resistant phenotype (Table 3). Additionally, four mutations were identified in genes such as *tet(A)*, *adeR*, *OprD*, and *gyrA*. Carbapenemases, including *bla*_OXA-23_, *bla*_OXA-113_, *bla*_OXA-174_, *bla*_OXA-312_, *bla*_OXA-72_ and the aminoglycoside acetyltransferase *aac(6’)-Ib7*, were predominantly found in the resistant category. The mutations, Abau_OprD_IPM|Asn411Asp, and Abau_gyrA_FLO|Ser81Leu were majorly found in the resistant category whereas tet(A)|Ala109Thr, and adeR|Leu142Ile, were found mainly in the susceptible category (Table 3). From the feature selection approach, we could identify the key features in KP and AB which can do the meropenem resistant and susceptible class discrimination. We further explored the association among the identified features contributing to the AMR phenotype.

**Table 3:** Details of 10 features identified from SVM-based feature selection for AB.

| AB feature name | SVM-coefficient | Chi-square p-value | Resistant (%) | Susceptible (%) |
| --- | --- | --- | --- | --- |
| *tet*(A)\|Ala109Thr | -1.383 | 0.040549136 | 17(8.85%) | 30(16.39%) |
| *adeR*\|Leu142Ile | 1.383 | 0.00567416 | 24(12.50%) | 44(24.04%) |
| *Abau_OprD_IPM*\|Asn411Asp | 0.83 | 3.28864E-06 | 190(98.96%) | 157(85.79%) |
| *Abau_gyrA_FLO*\|Ser81Leu | 1.384 | 1.36714E-15 | 192(100%) | 129(70.49%) |
| *bla*<sub>OXA-23</sub> | 2 | 1.14259E-33 | 114(59.38%) | 2(1.09%) |
| *bla*<sub>OXA-113</sub> | 2 | 0.026027857 | 7(3.65%) | 0 |
| *bla*<sub>OXA-174</sub> | 1.999 | 0.00861081 | 9(4.69%) | 0 |
| *bla*<sub>OXA-312</sub> | 2 | 0.000559352 | 14(7.29%) | 0 |
| *bla*<sub>OXA-72</sub> | 2 | 6.24106E-05 | 18(9.38%) | 0 |
| *aac*(6')-Ib7 | 1.66 | 0.001262163 | 15(7.81%) | 1(0.55%) |

### Association rule mining of the selected features

To uncover potential hidden associations among the identified features of KP and AB, we performed association rule mining using seven selected features for KP and 10 selected features for AB. For KP, we identified 23 rules with a support value of 0.01 and a confidence value of 0.80 (Table S7). To identify the factors associated with meropenem resistance in *K*P, we filtered the rules to focus on those with a meropenem-resistant phenotype as the consequent. We obtained five association rules that are linked to the resistant phenotype with a lift greater than two, involving four features of the AMR genes *bla*_KPC-2_, *bla*_KPC-3_, and *ble*_MBL_, *aac(6’)-Ib9*. The network representation of the five association rules contributing to the meropenem-resistant phenotype depicted in Fig. 5a indicates that the four AMR genes are associated with the meropenem-resistant phenotype (Table S7). Association rules indicate that the presence of any one of *bla*_KPC-2_, *bla*_KPC-3_, *ble*_MBL_ or *aac(6’)-Ib9* can individually contribute to meropenem resistance. The *aac(6’)-Ib9* gene is also found in combination with *bla*_KPC-3_ contributing to meropenem resistance (Table S7). We also identified seven rules involving mutations that contribute either individually or in combination to the meropenem-susceptible phenotype, with lift values ranging from 1.3 to 1.5. The three mutations were found to be in efflux pump genes.

**Fig. 5:**
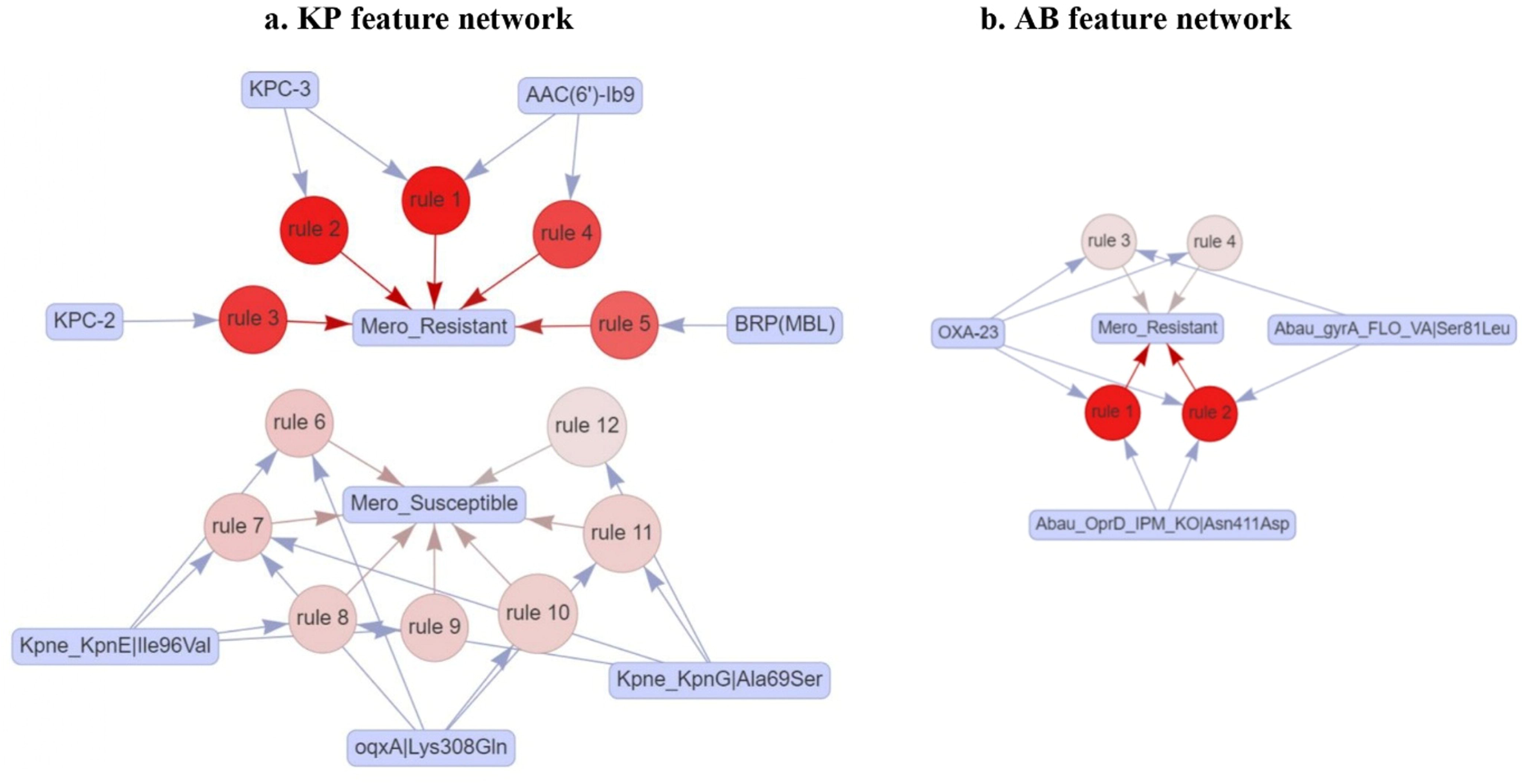
Network of 12 association rules based on AMR genes and mutations in KP (antecedents) contributing to meropenem resistance and susceptible (consequents). **b** Network of 4 rules based on AMR genes and mutations in AB(antecedents) contributing to meropenem resistance. The red shading indicates lift values ranging from 1.3 to 2.5 in KP and 1.91 to 1.93 in AB.

For AB, we identified 84 rules with a support value of 0.06 and a confidence value of 0.80 (Table S8). To pinpoint the factors contributing to meropenem resistance in AB, we filtered the rules to focus on those with a meropenem-resistant phenotype as the consequent, resulting in only four rules associated with the resistant phenotype. Among the 10 features used for association rule mining, three were found to be linked to the meropenem-resistant phenotype. These include one AMR gene (*bla*_OXA-23_) and two mutations (Abau_OprD_IPM|Asn411Asp, Abau_gyrA_FLO|Ser81Leu) with a lift of 1.91 to 1.93. The network representation of the association rules is shown in Fig. 5b. *bla*_OXA-23_ contributes individually to the meropenem-resistant phenotype. None of the mutations were individually contributed to the resistant phenotype; they contributed to resistance in combination with *bla*_OXA-23_ (Table S8).

### Analysis of antibiotic co-resistance and shared resistance patterns in KP and AB isolates

The isolates of KP and AB used in this study were found to be multi-drug resistant. Consequently, their genomes are expected to harbor features contributing to resistance against various antibiotics. To identify meropenem-associated co-resistance patterns with other antibiotics, we performed association rule mining using antibiotic resistance data available for the isolates. In KP, 24 different antibiotics from 9 classes were analysed. Using association rule mining with a support of 0.1, a lift value greater than 2 and a confidence value greater than 0.98, we identified 87 rules. Based on the analysis of antibiotic co-resistance, 18 antibiotics (ampicillin, ampicillin/sulbactam, aztreonam, cefazolin, cefepime, cefoxitin, ceftazidime, ceftriaxone, cefuroxime, ciprofloxacin, ertapenem, gentamicin, imipenem, levofloxacin, meropenem, piperacillin/tazobactam, tobramycin, and trimethoprim/sulfamethoxazole) from 7 different classes were found to be associated. The associated antibiotic classes in KP include carbapenems, cephalosporins, fluoroquinolones, monobactams, penicillins, aminoglycosides and combination antibiotics (Fig. 6a).

**Fig 6:**
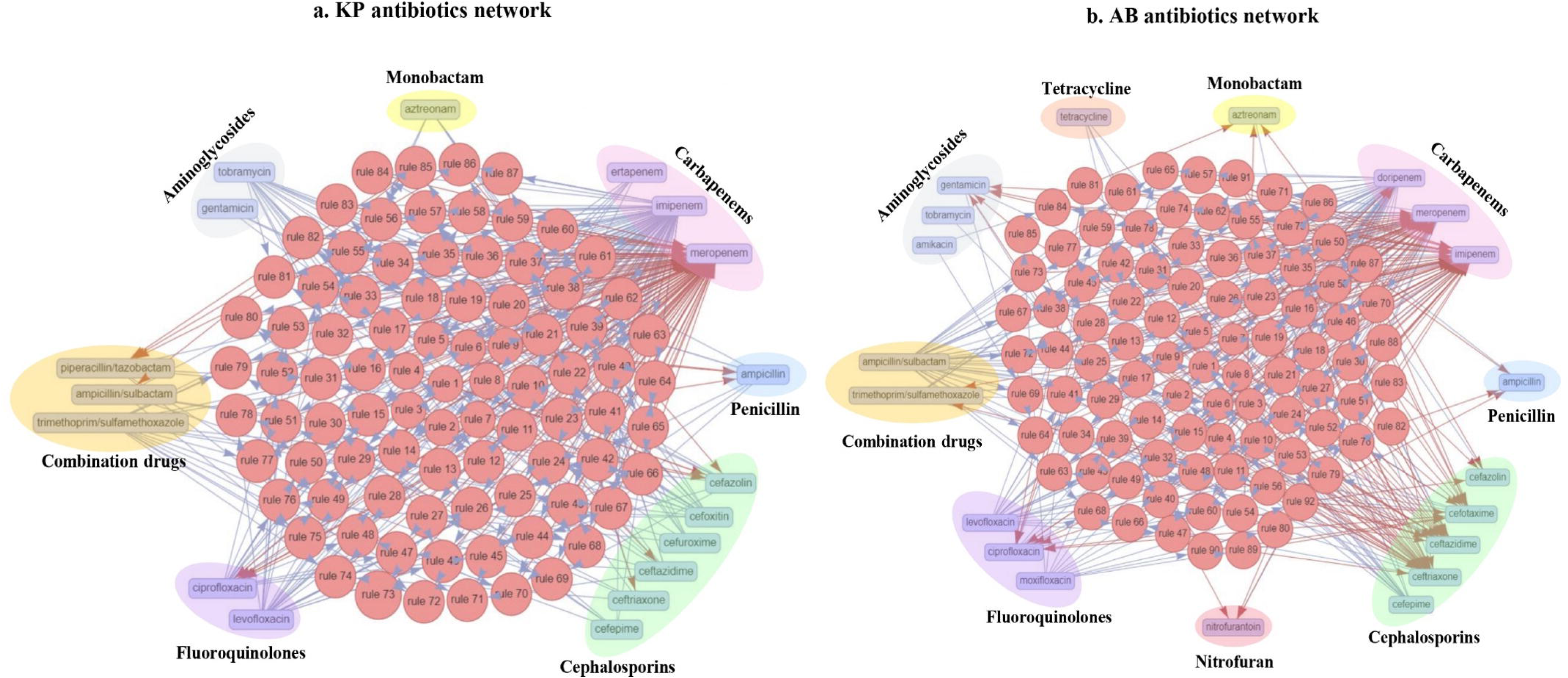
Network representation of shared and co-resistance among antibiotic classes. **a** Depicts 87 association rules in KP. **b** Depicts 92 association rules in AB.

In AB, 24 antibiotics from 10 different classes were included in the study. Using association rule mining with a support of 0.1, a lift value greater than 2 and a confidence value greater than 0.98, we identified 92 rules. Based on the analysis of antibiotic co-resistance, 20 antibiotics (imipenem, meropenem, doripenem, ceftazidime, trimethoprim/sulfamethoxazole, ampicillin, ceftriaxone, moxifloxacin, cefepime, ampicillin/sulbactam, levofloxacin, nitrofurantoin, tetracycline, cefotaxime, ciprofloxacin, gentamicin, cefazolin, aztreonam, tobramycin, amikacin) from 9 different classes were found to be associated. The antibiotic classes associated with AB include carbapenems, cephalosporins, combination antibiotics, penicillin, fluoroquinolones, nitrofuran, tetracyclines, aminoglycosides, and monobactam (Fig. 6b).

## Discussion

The broad-spectrum carbapenem antibiotic, meropenem, plays a critical role in treating severe infections caused by multidrug-resistant gram-negative bacteria. The emergence and spread of meropenem-resistant strains have significantly compromised their effectiveness, presenting a major threat to clinical practice. Understanding meropenem resistance is crucial, particularly from a global perspective, for developing effective strategies to preserve meropenem efficacy against infections. In this study, we employed large-scale global genome analysis of two ESKAPE pathogens, KP and AB, to investigate the genetic and evolutionary mechanisms by which they acquire meropenem resistance.

The KP and AB isolates studied were found to be multi-drug resistant, with their genomes harboring multiple factors contributing to resistance to various antibiotics. Our study aimed to investigate the genetic determinants of meropenem resistance, with the analysis potentially revealing hidden patterns of co-resistance or shared resistance to multiple antibiotics. Carbapenem resistance can arise from multiple mechanisms such as the production of antibiotic-inactivating enzymes, alterations in penicillin-binding proteins, inactivation of porin channels, overproduction of efflux pumps, or a combination of these factors (8). We investigated these possibilities by analysing the presence of AMR genes and their missense mutations in clinical isolates of KP and AB that are resistant or susceptible to meropenem. The metadata features of the clinical isolates, including ST, isolation source, year, and country showed no distinct patterns differentiating resistant and susceptible strains (Fig. 1a and Fig.1b).

In this study, we utilized AMR genes from the CARD as our reference for AMR gene and mutation identification. This extensive database provides regularly updated coverage of diverse resistance genes and mechanisms across bacterial species, enabling robust detection of AMR determinants compared with the limitations of using a single reference genome. The antibiotic efflux category of AMR genes, along with their corresponding regulators, emerges as the predominant group of AMR genes in both KP and AB. The antibiotic inactivation category, which includes the carbapenemase class of enzymes, was notably enriched in both organisms, with higher levels in the resistant strains compared to the susceptible strains (Fig. 2a and Fig. 2b). The reduced permeability category which includes porin genes, is more in KP and less in AB. Meropenem targets the PBP gene, which is categorized under target alterations, among which *pbp3* is present in KP. With respect to mutations, antibiotic target alteration and antibiotic efflux category exhibit a high mutation frequency in KP, while efflux pumps and reduced permeability to the antibiotic category were highly mutated in AB (Fig. 2c and Fig.2d). These findings from global genome analysis reveal distinct organism-specific distributions of resistance genes and mutations, highlighting the variability in resistance mechanisms between these two pathogens (38).

Considering the mechanisms contributing to resistance to meropenem, carbapenemases which that can hydrolyse meropenem are crucial. Class B metallo-β-lactamases, along with serine carbapenemases from classes A and D, have been shown to hydrolyse various carbapenems, including meropenem (39,40). In our study, more carbapenemases (44 genes) were observed in AB than in KP (16 genes). The carbapenemases *bla*_KPC-2_ and *bla*_KPC-3_ are predominantly found in meropenem-resistant KP isolates, and this observation is consistent with reports from other studies (41,42). The *bla*_OXA-23_ enzyme is predominantly found in resistant AB isolates, as reported in the literature (43). No mutations in the carbapenemases were identified in this study. Approximately 7% of the meropenem-resistant KP isolates did not contain carbapenemases, whereas all resistant isolates of AB harbor carbapenemases. The analysis of the presence of AMR genes and mutations revealed no specific difference between carbapenemase samples and non-carbapenemase samples. The possibility of resistance to carbapenem in non-carbapenemase samples may be due to the combined effects of beta-lactamase genes and the porin, efflux and PBP mutations (44,45). A study performed in non-carbapenemase carbapenem-resistant isolates of *Pseudomonas aeruginosa* revealed that mutations in the efflux, porin and PBP genes affect the susceptibility to carbapenem antibiotics (46)

A total of 1,900 features, including AMR genes and mutations in KP, and 1,303 features in AB, were identified through large-scale genome analysis to better understand the resistance mechanisms to meropenem. We applied various ML and DL models to predict resistance to meropenem. With the SVM model with linear kernel yielding the best performance on the basis of the evaluation metrics, a sequential feature selection approach was applied to identify important features that can discriminate between meropenem resistant and susceptible phenotypes in KP and AB. Seven features were selected in KP and 10 features in AB, achieving testing and external validation accuracies of 94% and 94% and 92% and 92% in KP and AB, respectively (Fig. 4b and Fig. 4d). Follow-up association mining was also conducted to identify the associations among these features with meropenem resistant and susceptible phenotypes.

The features most strongly associated with the meropenem-resistant phenotype in KP include four AMR genes (*bla*_KPC-2_, *bla*_KPC-3_, *ble*_MBL_, and *aac(6’)-Ib9*) and those associated with the meropenem-susceptible phenotype include three mutations (oqxA|Lys308Gln, Kpne_KpnE|Ile96Val, and Kpne_KpnG|Ala69Ser). Our multi-level analytics approach highlighted the carbapenemases *bla*_KPC-2_ and *bla*_KPC-3_ in KP as significant features associated with meropenem resistance (39), demonstrating the model’s discriminative power in selecting relevant features. The other carbapenemases didn’t turn up in our analysis because of their low prevalence in the isolates included in the study. BRP (MBL), a bleomycin resistance protein encoded by the *ble*_MBL_ gene, was a metallo-beta-lactamase that lacks intrinsic carbapenemase activity (47). Dortet et al. reported that the *ble*_MBL_ gene was predominantly found in *bla*_NDM-1_ producing bacteria, where it was co-expressed with the *bla*_NDM-1_ gene under the control of the same promoter as part of the same operon (47). Consistent with this, *ble*_MBL_ has been observed to coexist with *bla*_NDM-1_ within the same operon in the KP isolates used in present the study. Additionally, in some isolates, *bla*_NDM-5_ was found in place of *bla*_NDM-1_ within the same operon. Given that *ble*_MBL_ is a common element across multiple *bla*_NDM_ variants with carbapenemase activity, it emerged as a more significant feature than individual *bla*_NDM_ variants during the feature selection process. *aac(6’)-Ib9* is an aminoglycoside-modifying enzyme belonging to the AG N-acetyltransferases (AACs) family (48). The modification of aminoglycoside antibiotics reduces their affinity for bacterial ribosomes, contributing to drug resistance (49). *aac(6′)-Ib* is a prevalent and clinically significant aminoglycoside-modifying enzyme, with approximately 50 known variants identified in various gram-negative species (50). To investigate the potential link between *aac(6’)-Ib9* and meropenem resistance, we analysed genomes harboring this gene. In our study, the AAC gene was found in 64.12% of the genome, with *aac(6’)-Ib9* specifically detected in 2.11% of KP isolates. The co-occurrence analysis of the *aac(6’)-Ib9* gene and carbapenemase genes revealed that at least one carbapenemase gene was present in genomes containing *aac(6’)-Ib9*. Because of its co-occurrence with carbapenemase genes, *aac(6’)-Ib9* has emerged as a key factor associated with meropenem resistance. Interestingly, the antibiotic association analysis for KP, as reported in Fig. 6a, revealed co-resistance between aminoglycosides and carbapenems (51).

The mutation features in KP identified through the SVM model are associated with a susceptible phenotype and are found in efflux pump genes, including oqxA|Lys308Gln, Kpne_KpnE|Ile96Val, and Kpne_KpnG|Ala69Ser. Efflux pumps play a crucial role in antibiotic exclusion, and function-altering mutations in these pumps can contribute to antibiotic susceptibility. The efflux pumps *oqxA*, *kpnE*, and *kpnG* have been reported to contribute to carbapenem export (52,53), and mutations in these pumps may impact meropenem susceptibility.

In AB, 10 features were identified by the SVM model to classify meropenem resistant and susceptible phenotypes. This includes six AMR genes (bla_OXA-23_, *bla*_OXA-113_, *bla*_OXA-174_, *bla*_OXA-312_, *bla*_OXA-72_, *aac(6’)-Ib7*) and four mutations (tet(A)|Ala109Thr, adeR|Leu142Ile, Abau_OprD_IPM|Asn411Asp, Abau_gyrA_FLO|Ser81Leu). The investigation of the associations among these features to the meropenem-resistance phenotype revealed three features which include one AMR gene (*bla*_OXA-23_) and two mutations (Abau_OprD_IPM|Asn411Asp, Abau_gyrA_FLO|Ser81Leu). *bla*_OXA-23_, a key carbapenemase identified in our study, is a major factor contributing to meropenem resistance in AB (54). Additionally, other *bla*_OXA_ variants, including *bla*_OXA-72_, *bla*_OXA-113_ (55), and *bla*_OXA-312_ (56), *bla*_OXA-174_ (57) are known to contribute to carbapenem resistance in AB. However, other *bla*_OXA_ types were rarely found in the AB meropenem-resistant isolates and did not emerge as significant factors.

As observed in KP, an aminoglycoside acetyltransferase gene variant, *aac(6’)-Ib7* was a feature of AB. In our study, the AAC gene was found in 41.33% of the genome, with *aac(6’)-Ib7* specifically detected in 4.26% of the AB isolates. Similar to KP, all the isolates that harbor *aac(6’)-Ib7* contain a minimum of one carbapenemase and the observed occurrence of this enzyme with meropenem resistance may be incidental, resulting from co-resistance to both meropenem and aminoglycosides Fig. 6b. One notable mutation in AB is Abau_OprD_IPM|Asn411Asp in the porin gene, which is involved in carbapenem diffusion (58), and is present in 92.53% of all isolates and 98.95% of meropenem-resistant isolates. *OprD* mutations are widely reported in multidrug-resistant AB, where they contribute to resistance mechanisms (59), but no specific details are available for this mutation. Another mutation identified was Abau_gyrA_FLO|Ser81Leu, which is present in almost 85.60% of all the isolates and was found in all the resistant isolates. Nancy M Attia et al. reported that Ser81Leu in *gyrA* led to fluoroquinolone resistance in AB. Our antibiotic association study revealed co-resistance among carbapenem and fluoroquinolone antibiotics (Fig. 6b) and the association of this feature with the meropenem-resistance phenotype in AB can be by chance due to the co-resistance to fluoroquinolone antibiotics (60).

The genetic determinant analysis of meropenem resistance revealed multiple antibiotic resistance factors, which led us to conduct an association study to explore their interrelationships across different antibiotics. The antibiotic classes include cephalosporins, carbapenems, fluoroquinolones, combination antibiotics (piperacillin/tazobactam, ampicillin/sulbactam, trimethoprim/sulfamethoxazole), penicillin, aminoglycosides, and monobactam (61). We observed co-resistance between carbapenem and aminoglycoside in both KP and AB contributed by AAC family genes and carbapenemases (62–64). Carbapenem and fluoroquinolone were found to be associated with AB contributed by the prevalence of gyrA mutations in carbapenem-resistant isolates. The porin and efflux pump gene mutations suggest the potential for shared resistance across multiple antibiotics

Large-scale genomic analysis using ML/DL techniques can effectively uncover genetic factors that effectively drive meropenem resistance and explore multiple antibiotic resistance relationships. The major observations from the present study are as follows: (1) Differences in AMR genes and mutations across various resistance mechanisms contribute to organism-specific resistance; (2) Carbapenemases as major factors, along with mutations in porin and efflux pumps contribute to meropenem resistance; and (3) Shared or co-resistance across antibiotic classes, with genetic factors linked to carbapenem, aminoglycoside, and fluoroquinolone resistance. However, there are some limitations. The sample size for AB is smaller than that for KP. Additionally, we focused only on the AMR genes from the CARD database and mutations within those genes. The results of the model parameters and optimization algorithms are closely linked to the dataset, and any changes to these parameters may affect the model’s features. Our observations suggest that gene presence alone does not confirm resistance, and a multi-omics profile is essential to uncover additional contributing factors. Given the inherent diversity of genetic determinants, including spontaneous mutations influenced by environmental factors and mobile genetic elements, pinpointing specific determinants of meropenem resistance across all global strains remains a challenge.

## Conclusion

Through large-scale genome analysis, this study highlights the genetic factors underpinning meropenem resistance in two major ESKAPE pathogens, KP and AB. Our exploratory data analysis of carbapenem resistance mechanisms revealed differences in the proportion of resistance factors, including carbapenemases, efflux pumps, porins, and antibiotic target alterations in KP and AB. The resistance mechanisms were similar in the meropenem resistant and susceptible genomes of KP and AB, with a slight increase in antibiotic inactivation in the resistant group. Using ML/DL and association mining approaches, we could underscore the significant role of AMR genes such as *bla*_KPC-2_, *bla*_KPC-3_, *ble*_MBL_, and *aac(6’)-Ib9* in KP, and *bla*_OXA-23_ in AB in meropenem resistance. Mutations in porin, efflux and target alteration categories are key features. Follow-up association mining revealed the combined effect of mutations with other features contributing to meropenem resistance. The presence of AAC genes and *gyrA* mutations in meropenem-resistant isolates indicates the co-resistance with aminoglycosides and fluoroquinolone antibiotics, and porin and efflux mutations indicate the possibility of shared resistance among antibiotics. A global analysis of antibiotic associations revealed co-resistance among multiple classes of antibiotics in KP and AB, highlighting the urgent need for enhanced resistance control and revised antibiotic usage policies to support SDG 3: Good Health and Well-being.

## DATA AVAILABILITY STATEMENT

The study used publicly available data from the BVBRC and NCBI Pathogen databases. The details of the dataset are provided in the supplementary files.

## AUTHOR CONTRIBUTIONS

SM & SR conceived, optimised the study design. DK, AS, and AV were analysed and performed scripting. SR, DK, AS, AV, SK, MR, DD, and SB performed data analysis. SR investigated and validated the study. SR & SM contributed to drafting and revising the manuscript. All authors have read and agreed to the final version of the manuscript.

## Data Availability

This study utilized publicly available clinical isolate bacterial genome data, with dataset details provided in the supplementary table.

## Supplementary Figures

**Figure S1:**
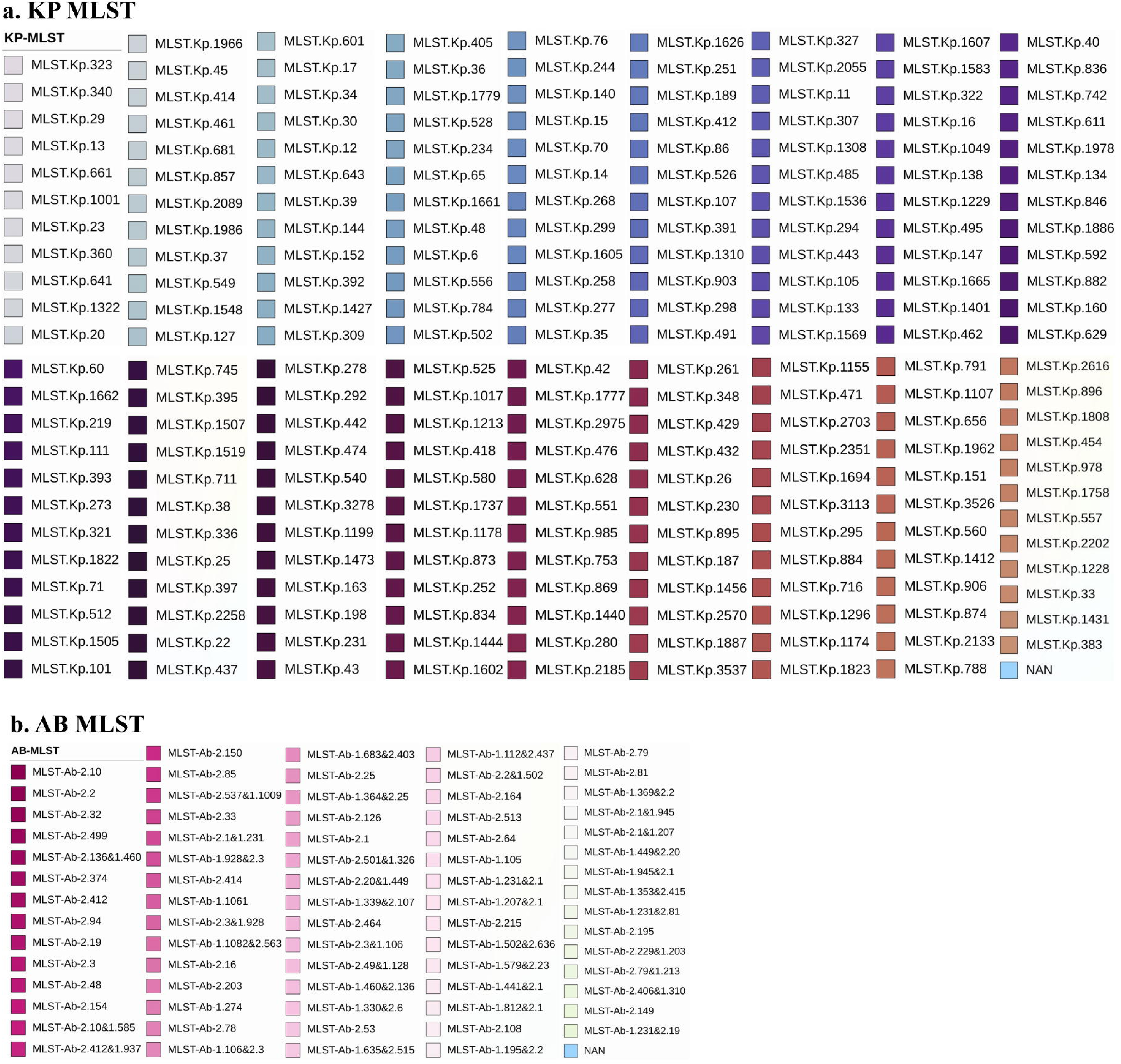
The phylogenetic tree legend includes 216 unique MLST types for **a** KP and 75 unique MLST types for **b** AB

**Figure S2:**
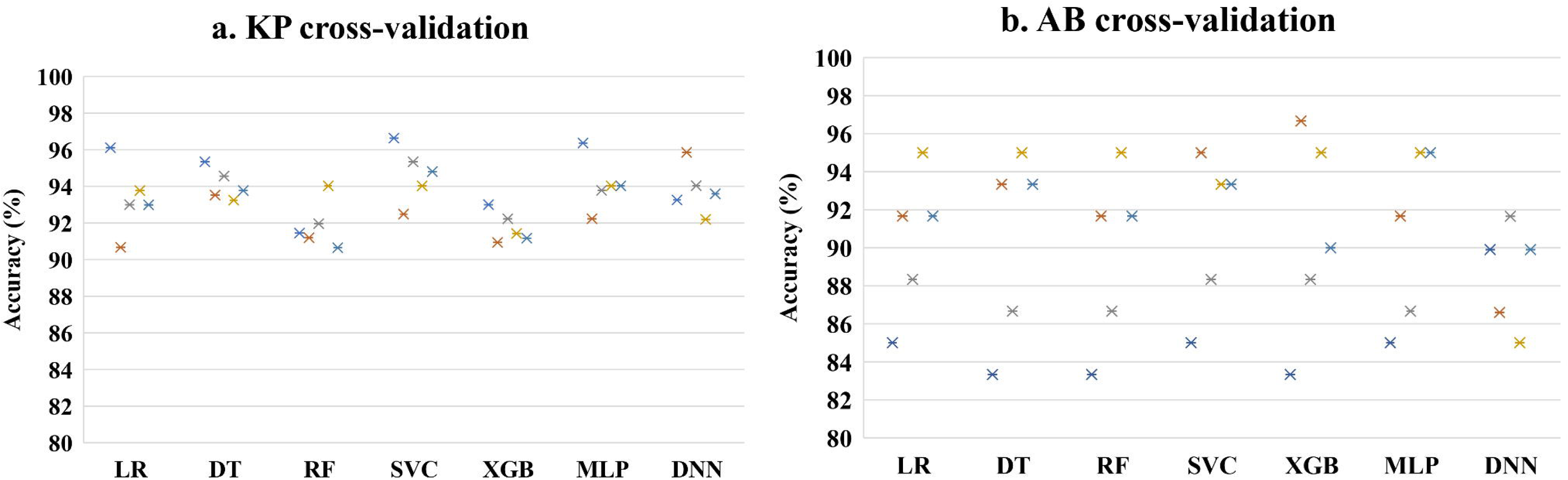
Five-fold cross-validation accuracy for six ML models and one DL model, optimized using Bayesian-based hyperparameter tuning for **a** KP and **b** AB

